# Association between LVEF-1 and Coronary Artery Stenosis and Interventional Treatment Efficacy

**DOI:** 10.1101/2024.12.30.24319795

**Authors:** Chao Tang, Bin Yan, Li Xiong, Yanyu Zhu, Jiaxing Ren, Xiang Li, Xiaosong Gu

## Abstract

**Background:** Myocardial ischemia leads to a decrease in the speed and intensity of myocardial tissue contraction. In the early stages, this cardiac dysfunction is difficult to be detected by conventional echocardiography methods.

**Objective:** First-phase left ventricular ejection fraction (LVEF-1) serves as a sensitive indicator for evaluating myocardial contractility. The aim of this study was to assess the decreased cardiac function caused by myocardial ischemia and its improvement following interventional treatment, as measured by LVEF-1.

**Methods:** 207 patients were enrolled. Based on the results of the angiography, they were categorized into three groups: mild, moderate, and severe. The LVEF-1, Gensini score and other clinical data were compared among these groups. Furthermore, logistic regression analysis was used to analyze the relationship between LVEF-1 and coronary artery stenosis, as well as factors associated with improvement in LVEF-1 following interventional treatment using linear regression.

**Results:** The LVEF-1 of patients in the 3 groups were 29.6 (28.2, 31.7) %, 27.8 (27.0, 28.6) %, and 25.2 (23.6, 26.5) % (p<0.001). There was a negative relationship between LVEF-1 and the Gensini score (r=-0.694, p<0.001), as well as between LVEF-1 and BNP (r=-0.244, p<0.001). LVEF-1 was identified as an independent predictor for coronary artery stenosis ≥50% or ≥70%. A cutoff value of 26.9% for LVEF-1 had a sensitivity of 89.5% and specificity of 83.9% for predicting the coronary artery stenosis ≥70%. Following intervention, LVEF-1 increased from 24.70 (23.30, 26.32) % to 28.10 (26.80, 29.92) % in 82 patients. Stent diameter was identified as an independent factor influencing the improvement in LVEF-1 post-intervention.

**Conclusions:** LVEF-1 is negatively correlated with the severity of coronary artery stenosis, and it increases after receiving coronary artery intervention therapy, suggesting that LVEF-1 can serve as a new indicator to evaluate the severity of coronary artery stenosis and the efficacy of interventional treatment.

## Introduction

In clinical practice, many patients with coronary heart disease (CHD) do not exhibit any difference in left ventricular ejection fraction (LVEF) compared to normal individuals. A significant decrease in LVEF can only be detected in cases of acute myocardial infarction or chronic ischemic cardiomyopathy. Early changes in cardiac function in CHD patients are difficult to detect, and there is also a lack of effective evaluation methods for rapidly assessing the improvement of cardiac function after revascularization through percutaneous coronary intervention (PCI).

Echocardiography plays an important role in clinical practice for assessing and aiding in the diagnosis of CHD. However, traditional echocardiographic parameters such as LVEF and the detection of segmental wall motion abnormalities may not always reveal significant abnormalities in patients with partially occluded coronary arteries or small myocardial infarctions compensating in other areas. New technologies, such as two-dimensional speckle tracking echocardiography and three-dimensional echocardiography, have been developed to address these limitations ^[1]^. Myocardial perfusion is also a valuable tool for evaluating myocardial function ^[2, 3]^. Nevertheless, these methods involve complex examination and analysis procedures, often requiring specialized software for post-processing of the original images, which can impede their widespread adoption in clinical settings. Therefore, there is a pressing desire to explore a briefer echocardiographic index that can efficiently and precisely identify cardiac dysfunction in CHD patients, as well as improve the evaluation of myocardial recovery following revascularization in these individuals.

First-phase left ventricular ejection fraction (LVEF-1) is an innovative echocardiographic index proposed by Gu Haotian et al. in 2017 ^[4]^. This index refers to the proportion of left ventricular volume change during the time from aortic valve opening to the peak aortic blood flow velocity, as a percentage of the end-diastolic left ventricular volume. It can be quantified using transthoracic echocardiography. In recent years, many studies have found that LVEF-1 is strongly correlated with left ventricular systolic and diastolic functions, being sensitive to early changes in heart disease ^[4, 5]^. Therefore, there is a possibility of regarding LVEF-1 as a novel indicator for assessing myocardial ischemia. Indeed, Minczykowski et al. have found that patients with CHD are more likely to have lower LVEF-1 by comparing parameters from healthy individuals and patients with CHD ^[6]^. However, they did not further investigate the association between LVEF-1 and the severity of coronary artery stenosis. Therefore, we designed this study to investigate the association between LVEF-1 and the degree of coronary artery stenosis, as well as any changes in LVEF-1 pre and post PCI.

## Methods and Materials

### 1. Study population

Patients who underwent coronary angiography in the Jingjiang people’s hospital and the Second Affiliated Hospital of Soochow University for the first time due to chest pain between June 2021 and June 2023 were consecutively enrolled in the study (Figure 1). Inclusion criteria were: 1. Age ≥ 18 years; 2. Presence of symptoms of angina or clinical evidence of myocardial ischemia; 3. Preoperative echocardiography showing a LVEF greater than 50%; 4. Willingness to participate. Exclusion criteria were: 1. History of myocardial infarction (acute or chronic) or heart failure, LVEF < 50%; 2. Presence of valvular heart disease (including moderate or severe valve regurgitation, and any degree of valve stenosis), malignant arrhythmias, atrial fibrillation, or congenital heart disease; 3. Chronic total occlusion of coronary artery; 4. Poor baseline conditions such as liver or kidney dysfunction, tumors, or infections; 5. Poor quality echocardiography images.

**Figure 1.**
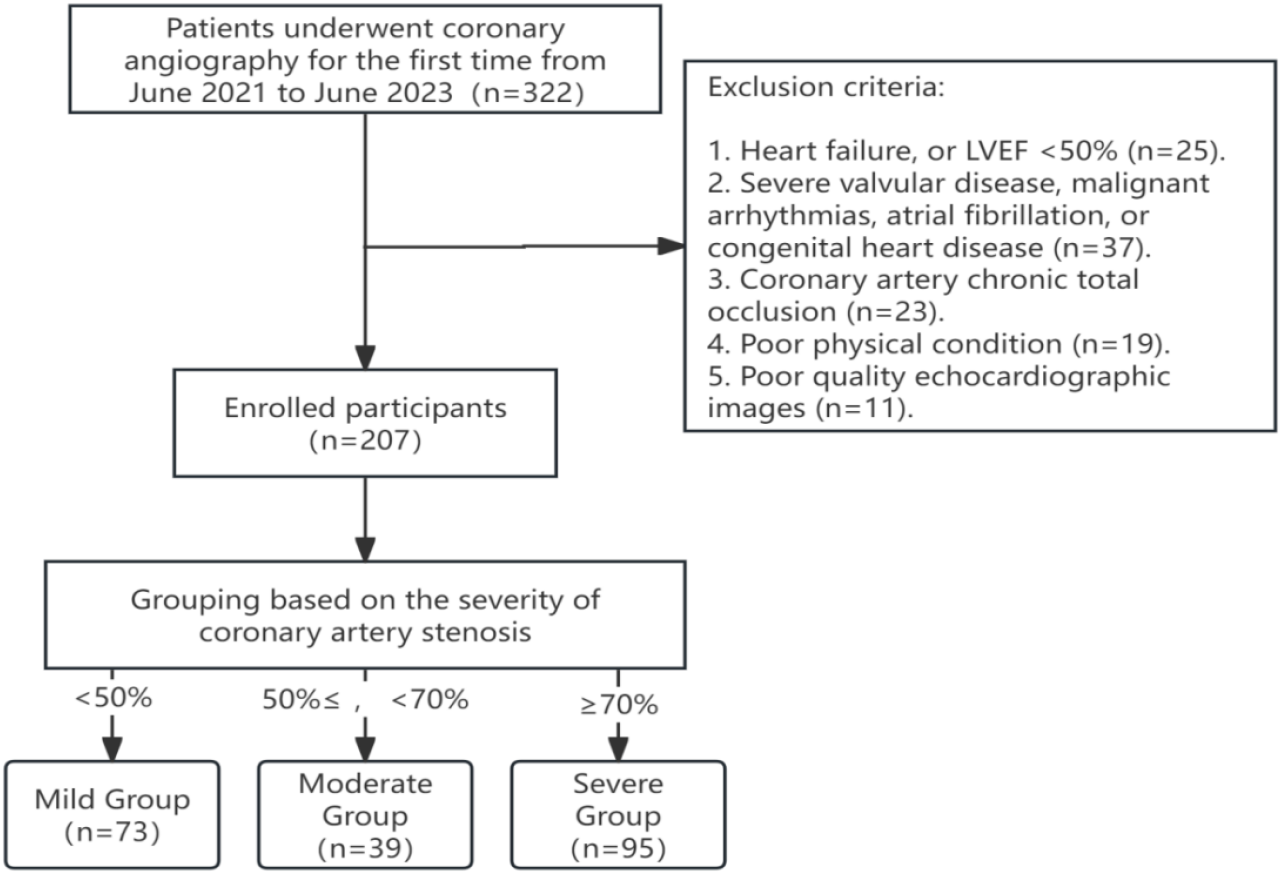
Flowchart of patients enrollment process

The study protocol was developed in line with the declaration of Helsinki. Informed consent was obtained from all participants. This study was approved by the Ethics Committee of Jingjiang People’s Hospital (2022-KY-014-01).

### 2. General data

Clinical data: including age, gender, Body Mass Index (BMI), blood pressure (including both systolic blood pressure (SBP) and diastolic blood pressure (DBP), heart rate (HR), and history of smoking, hypertension (HBP), and diabetes mellitus (DM).

Blood test results: fasting venous blood samples were collected from all patients and analyzed by the hospital laboratory. The data collected included hemoglobin (Hb), glucose (Glu), total cholesterol (TC), triglycerides (TG), low-density lipoprotein (LDL), high-density lipoprotein (HDL), creatinine (CREA), C-reactive protein (CRP), and B-type natriuretic peptide (BNP).

Coronary angiography: Following coronary angiography, patients were classified based on the degree of maximal stenosis: mild group (coronary artery maximal stenosis <50%), moderate group (50%≤ coronary artery maximal stenosis <70%), and severe group (coronary artery maximal stenosis ≥70%). The Gensini score was then calculated ^[7]^. For patients who underwent PCI, the diameter and length of stents used were recorded.

### 3. Echocardiographic parameters

All echocardiographic examinations were performed by a professional physician using a Philips EPIQ 7C echocardiography system on the day of coronary angiography or 1-2 days before the procedure. The physicians were blinded to the patients’ clinical data. Patients undergoing PCI treatment received a repeat echocardiographic examination on the 4th day postoperatively before discharge. Each examination recorded left atrial diameter (LAD), left ventricular internal dimension diastole (LVDd), left ventricular internal dimension systole (LVDs), LVEF, global longitudinal peak systolic strain (GLS, Supplementary Figure 1) measured using speckle tracking echocardiography, and LVEF-1. Additionally, the early diastolic peak velocity of the mitral valve (E) and the early diastolic peak velocity of the lateral mitral annulus (e’) were measured, and E/e’ was calculated.

LVEF-1 measurement (Supplementary Figure 2): The continuous wave (CW) Doppler was used to obtain the aortic valve flow spectrum, measuring the time T1 from aortic valve opening to the peak aortic valve flow velocity. The apical four-chamber and apical two-chamber views was achieved, LV end-diastolic volume (LVEDV), LV end-systolic volume (LVESV), and the left ventricular volume at time T1 (T1V) were measured. LVEF and LVEF-1 were then calculated using the formulas LVEF= (LVEDV - LVESV)/LVEDV ×100% and LVEF-1= (LVEDV - T1V)/LVEDV ×100% ^[4]^.

**Figure 2.**
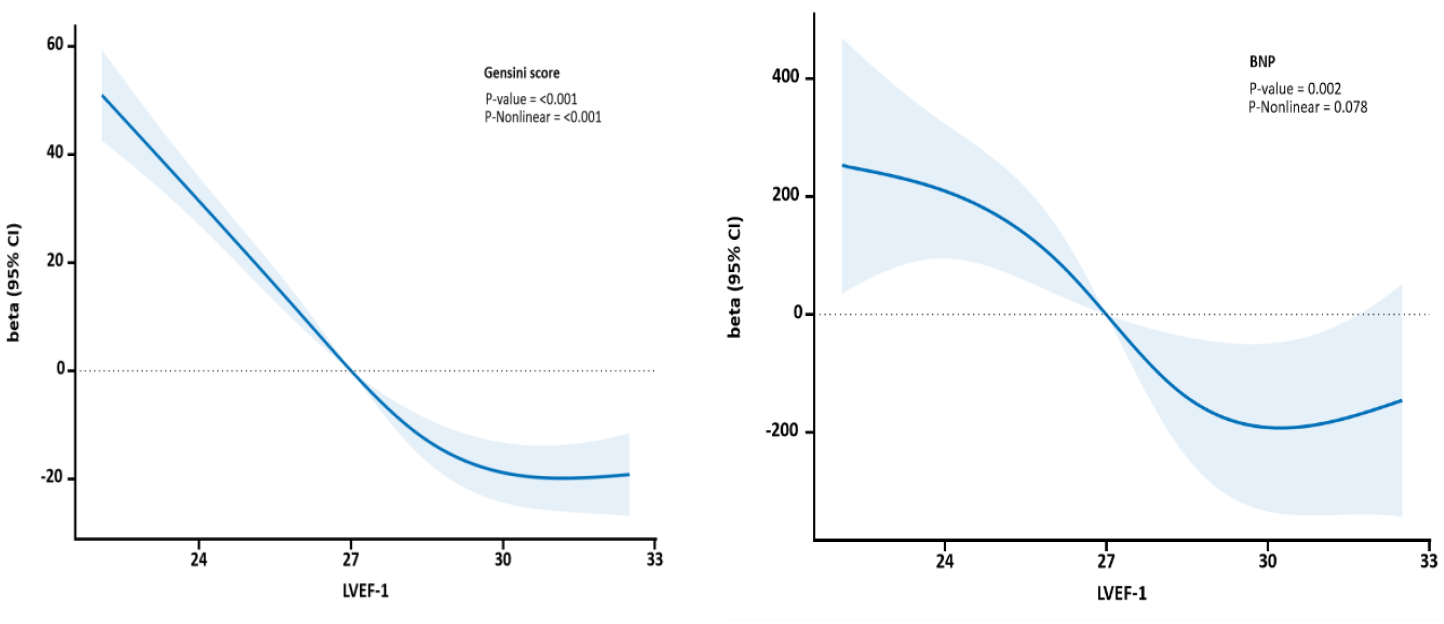
Association between LVEF-1 and Gensini score and BNP with the RCS function. Both models are with 4 knots. Y-axis represents the beta to present Gensini score for any value of LVEF-1 compared to individuals with reference value (50th percentile) of LVEF-1.

### 4. Statistical Methods

All statistical analyses were performed using SPSS and R, with the main R package “rms” utilized. Normally distributed data were expressed as mean ± standard deviation, and intergroup comparisons were conducted using either ANOVA or t-tests. Non-normally distributed data were expressed as median (interquartile range), and intergroup comparisons were conducted using the Kruskal-Wallis test. For categorical or ordinal data, frequencies (%) were reported, and intergroup comparisons were performed using the chi-square test.

Pearson’s correlation is used to test the relationship between variables, and restricted cubic splines (RCS) analysis was used to analyze the quantitative relationship between LVEF-1 and the severity of coronary artery stenosis and BNP. Univariate and multivariate logistic regression analyses were conducted to analyze the relationship between LVEF-1 and moderate or severe coronary artery stenosis. Paired t-tests were used to compare patient data before and after PCI. Multiple linear regression analysis was employed to investigate factors influencing the pre- and postoperative changes in LVEF-1. Receiver-operating characteristic (ROC) curve analysis and the respective area under the curve (AUC) were utilized to evaluate the diagnostic value of different indicators for coronary artery stenosis.

## Results

### 1. Baseline data comparison

According the results of coronary angiography, there were 39 patients in the moderate stenosis group, 73 patients in the mild stenosis group and 95 patients in the severe stenosis group. In the severe stenosis group, 82 patients underwent PCI treatment, while 12 patients did not undergo PCI treatment due to economic or other reasons. As the degree of stenosis increased, the proportion of male patients, history of smoking, HBP and DM all increased (P < 0.05). Additionally, SBP, DBP, Glu, CRP, BNP, LAD, LVDd, LVDs, E/e’, and Gensini score all showed an increasing trend (P < 0.05), while GLS, and LVEF-1 decreased (P < 0.05). LVEF among the three groups has no statistically significant, (Table 1).

**Table 1:**
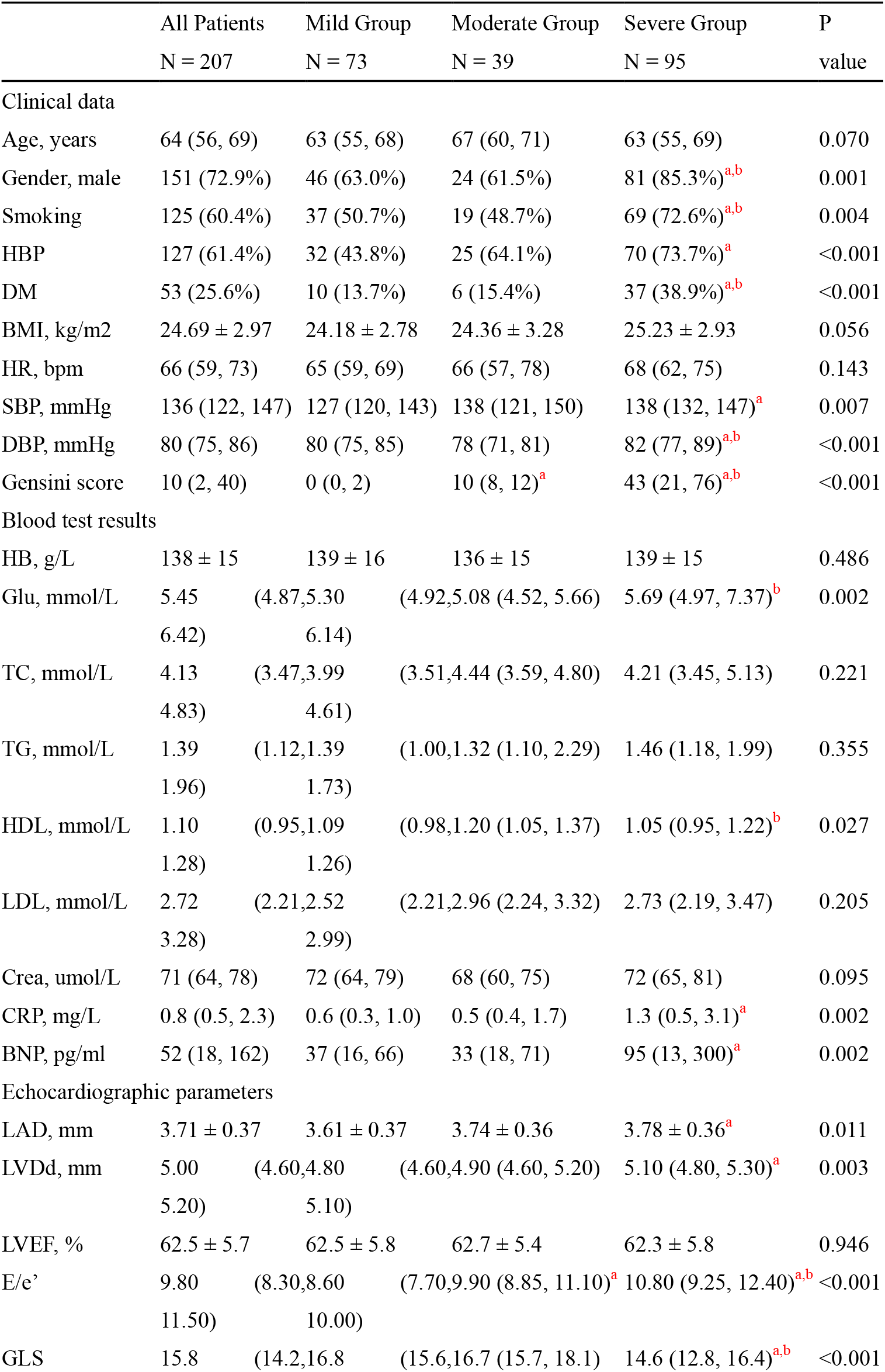
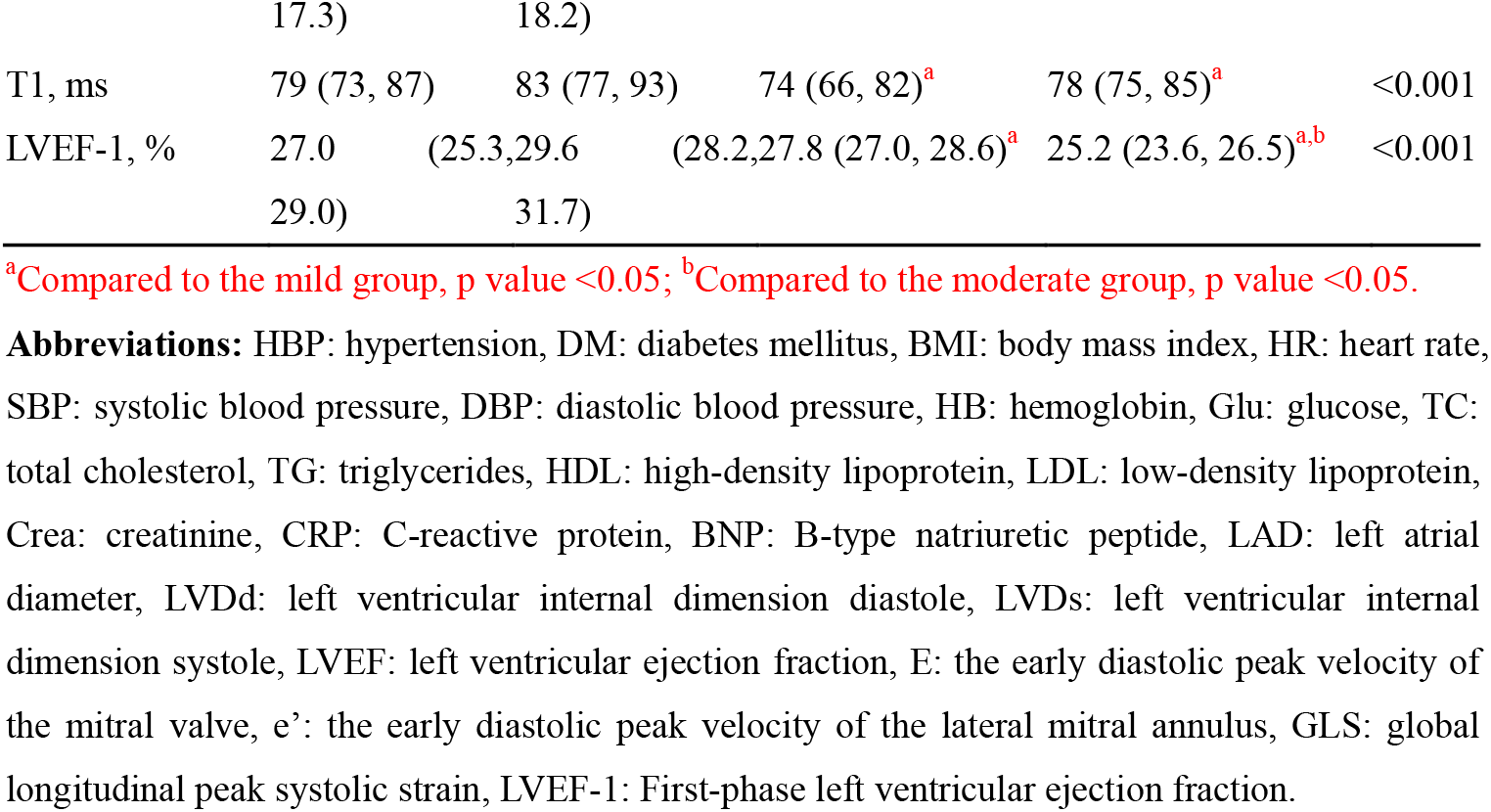
Comparison of data on coronary artery stenosis severity.

### 1. Correlation Analysis of LVEF-1 with clinical parameters

Correlation analysis was conducted between LVEF-1 and patients’ parameters (Supplementary Table 1). Gender, smoking, SBP, DBP, Glu, and LDL are negatively correlated with LVEF-1. Specifically, LVEF-1 showed a relatively negative correlation with the Gensini score (r= -0.694, P<0.01) and BNP (r= -0.244, P<0.01). Furthermore, RCS analysis indicated a non-linear negative correlation between LVEF-1 and the Gensini score (P < 0.001, and P_Nonlinear value_ < 0.001), as well as a linear negative correlation between LVEF-1 and BNP (P = 0.002, and P_Nonlinear value_ = 0.078). (Figure 2)

### 3. Value of LVEF-1 in the Assessment of Moderate or Severe Coronary Artery Stenosis

Univariate logistic regression revealed that LVEF-1 had a statistically significant predictive value for coronary artery stenosis ≥50% (OR=0.406, 95%CI 0.313-0.526, P<0.001) and coronary artery stenosis ≥70% (OR=0.366, 95%CI 0.275-0.486, P<0.001). This significance remained evident in various multivariable logistic regression models after adjusting for gender, age, smoking, HBP, DM, Glu, TG, HDL, LDL, CREA, and CRP (Table 2).

**Table 2:** Logistic regression analysis of LVEF-1 with coronary artery stenosis ≥50% or ≥70%.

| | Coronary artery stenosis $\geq 50\%$ | | Coronary artery stenosis $\geq 70\%$ | |
| --- | --- | --- | --- | --- |
|  | OR (95% CI) | P value | OR (95% CI) | P value |
| Univariate | 0.406 (0.313-0.526) | $<0.001$ | 0.366 (0.275-0.486) | $<0.001$ |
| Multivariate |  |  |  |  |
| model1 | 0.414 (0.318-0.538) | $<0.001$ | 0.352 (0.260-0.478) | $<0.001$ |
| model2 | 0.425 (0.325-0.556) | $<0.001$ | 0.373 (0.276-0.505) | $<0.001$ |
| model3 | 0.392 (0.290-0.530) | $<0.001$ | 0.355 (0.253-0.497) | $<0.001$ |

Model1: adjusted for gender and age
Model2: adjusted for model 1 + smoking, HBP, DM and BMI
Model3: adjusted for model 2 + Glu, TG, HDL, LDL, CREA and CRP
**Abbreviations:** HBP: hypertension, DM: diabetes mellitus, BMI: body mass index, Glu: glucose, TG: triglycerides, HDL: high-density lipoprotein, LDL: low-density lipoprotein, Crea: creatinine, CRP: C-reactive protein.

The ROC analysis compares LVEF-1, GLS, E/e’, LVEF, and LVDd for predicting coronary artery stenosis ≥50% or ≥70%. The AUCs were highest for LVEF-1 and significantly greater than that for other variables (P<0.001). A cutoff value of 27.9% for LVEF-1 had a sensitivity of 82.8% and specificity of 83.6% for predicting the coronary artery stenosis ≥50%. Additionally, a cutoff value of 26.9% for LVEF-1 had a sensitivity of 89.5% and specificity of 83.9% for predicting the coronary artery stenosis ≥70% (Figure 3).

**Figure 3.**
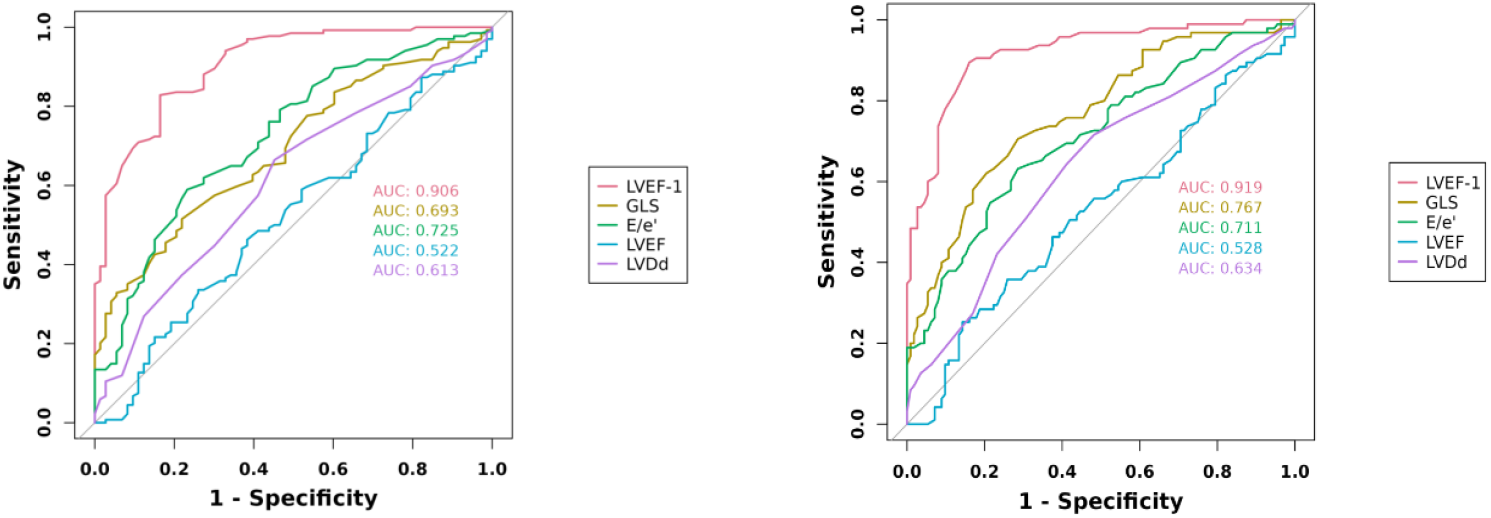
a ROC analysis compares different echocardiographic parameters for predicting coronary artery stenosis ≥50%. b ROC analysis compares different echocardiographic parameters for predicting coronary artery stenosis ≥70%.

### 4. Comparison of Echocardiographic Parameters in Patients Before and After PCI

During a comparison of echocardiographic parameters in 82 patients before and after undergoing PCI treatment, it was observed that following PCI, the E/e’ ratio decreased compared to pre-operation, while GLS and LVEF-1 both increased (P<0.05), (Table 3).

**Table 3:** Comparison of Echocardiographic Parameters in Patients Before and After PCI.

|  | Before PCI | After PCI | P value |
| --- | --- | --- | --- |
| HR, bpm | 68 (62, 75) | 68 (63, 73) | 0.925 |
| SBP, mmHg | 138(132,148) | 120(113,130) | <0.001 |
| DBP, mmHg | 83(77,89) | 70(65,76) | <0.001 |
| LAD, mm | 3.77±0.35 | 3.81±0.31 | 0.107 |
| LVDd, mm | 5.00(4.78,5.22) | 5.00(4.80,5.22) | 0.109 |
| LVDs, mm | 3.40(3.10,3.62) | 3.40(3.20,3.62) | 0.316 |
| LVEF, % | 59.2 ± 6.0 | 59.3 ± 5.5 | 0.705 |
| E/e' | 11.10(9.45,12.70) | 9.90(8.60,12.18) | <0.001 |
| GLS | 14.35(12.50,15.72) | 15.00(12.90,16.12) | 0.031 |
| T1, ms | 78(76,86) | 79(75,83) | 0.385 |
| LVEF-1, % | 24.70 (23.30, 26.32) | 28.10 (26.80, 29.92) | <0.001 |

**Abbreviations:** HR: heart rate, SBP: systolic blood pressure, DBP: diastolic blood pressure, LAD: left atrial diameter, LVDd: left ventricular internal dimension diastole, LVDs: left ventricular internal dimension systole, LVEF: left ventricular ejection fraction, E: the early diastolic peak velocity of the mitral valve, e': the early diastolic peak velocity of the lateral mitral annulus, GLS: global longitudinal peak systolic strain, LVEF-1: First-phase left ventricular ejection fraction

### 5. Multivariate Linear Regression Analysis of Factors Influencing Changes in LVEF-1 in Patients Before and After PCI

ΔLVEF-1 represents changes in LVEF-1 of the patients before and after PCI. Correlation analysis was performed between the clinical data in Table 1, postoperative SBP, DBP, changes in SBP and DBP, and ΔLVEF-1. The results indicated that age, Gensini score, and stent diameter had a p-value of less than 0.10 with respect to ΔLVEF-1. These three factors were included in the multivariate linear regression analysis. The results showed that Gensini score and stent diameter were independent factors influencing ΔLVEF-1, (Table 4).

**Table 4:** Multivariate linear regression analysis of factors influencing changes in LVEF-1 in patients before and after PCI.

|  | Estimate | SE | 95% CI Lower | 95% CI Upper | t | p-value |
| --- | --- | --- | --- | --- | --- | --- |
| Age, years | -0.023 | 0.014 | -0.050 | 0.003 | -1.739 | 0.086 |
| Gensini score | 0.012 | 0.004 | 0.003 | 0.021 | 2.745 | 0.008 |
| Diameter of stent, mm | 3.232 | 0.351 | 2.534 | 3.930 | 9.217 | <0.001 |

## Discussion

In our study, LVEF-1 showed a significant decrease in the group with severe coronary stenosis and demonstrated a negative correlation with the severity of coronary artery stenosis. Specifically, LVEF-1 was identified as an independent predictive factor for moderate and severe coronary artery stenosis. Following coronary intervention, there was a significant increase in LVEF-1 levels. It was found that the diameter of the cardiac stent(s), rather than the total length, was a factor that influenced the improvement of LVEF-1 after PCI.

The systolic function of the LV is a complex process. In the early stages of impaired LV systolic function, length-dependent activation of myocardial tissue may result in slower but sustained contraction to preserve ejection fraction ^[8]^. This leads to a situation where traditional measures of “systolic function” such as LVEF may be inadequate in detecting early systolic dysfunction in patients with CHD. More advanced echocardiographic indices, such as two-dimensional speckle tracking imaging (2D-STI), real-time three-dimensional echocardiography (RT-3DE), and three-dimensional speckle tracking imaging (3D-STI), are useful ^[9-11]^. However, these tests are technically complex, time-consuming, and difficult to implement on a large scale in clinical practice. Our study shows that within different stenosis groups, there is no difference in LVEF, while LVEF-1, as a simpler echocardiographic index, is strongly correlated with the severity of coronary artery stenosis. These results indicate that LVEF-1, the speed-dependent contraction of myocardial tissue, is a more sensitive marker than LVEF, the length-dependent activation of myocardial tissue, to assess early changes in myocardial ischemia. Additionally, our study found that the value of LVEF-1 in assessing moderate and severe stenosis in the coronary arteries is higher than GLS obtained based on 2D-STI, with a higher AUC, although GLS also has independent value in evaluating moderate and severe stenosis in the coronary arteries (Supplementary Table 2).

Currently, researches on LVEF-1 mainly focus on aortic valve stenosis and hypertension, indicating that increased cardiac afterload, as represented by these two conditions, can reduce early cardiac systolic function, resulting in a decrease in LVEF-1 ^[4,12-15]^. However, studies have also shown that in healthy individuals, after marathon running or appropriate exercise, there is an increase in LVEF-1 due to increased cardiac afterload ^[16,17]^. This seems to contradict the idea that increased afterload decreases LVEF-1. It is important to note that these studies focused on healthy individuals, and the increase in afterload was temporary. This may be related to the fact that a healthy heart still has a good contractile reserve function when dealing with sudden pressure. In contrast, sustained afterload increase due to aortic stenosis or hypertension undoubtedly weakens cardiac reserve. Our study found that higher blood pressure has a negative impact on LVEF-1, but in the short term after PCI, the increase in LVEF-1 seems to have little correlation with blood pressure and its changes. This suggests that the relationship between blood pressure and LVEF-1 in CHD patients may be more complex.

As we all know, myocardial ischemia is responsible for decreasing myocardial contractility ^[18, 19]^. LVEF-1 is a marker of myocardial contractility in ultrasound examination and reflects early changes in cardiac function under ischemic conditions, as demonstrated in our study. Previous research on stable CHD patients also shows that the LVEF in CHD patients is similar to that in healthy individuals, but their LVEF-1 is significantly lower ^[6]^. As suggested by Gu et al., to maintain global LVEF, myocardial contraction may last longer and sometimes extend beyond the duration of systole, leading to wasted and less effective myocardial work ^[4]^. This phenomenon is likely due to more severe coronary artery stenosis leading to increased myocardial ischemia, which in turn decreases myocardial work efficiency. Our study also finds that LVEF-1 has a relatively strong negative correlation with the Gensini score, and after PCI, LVEF-1 significantly increases. This result suggests that a higher LVEF-1 may correspond to better myocardial work efficiency in CHD patients.

We also found that in patients with coronary artery stenosis, LVEF-1 is negatively correlated with certain risk factors for CHD, such as male gender, high blood pressure levels, glucose, and LDL. Therefore, it is reasonable to speculate that even if coronary artery stenosis has not progressed to a severe degree, these risk factors may still contribute to impaired cardiac function in the early stages. Additionally, there is a significant negative correlation between LVEF-1 and BNP. Previous studies have also confirmed the correlation between BNP and the severity of coronary artery stenosis, with BNP being closely related to cardiac function and prognosis in patients with CHD ^[20,21]^. The association between LVEF-1 and BNP further confirms the importance of LVEF-1 in assessing cardiac function in patients with coronary artery stenosis.

LVEF-1 is not only independently associated with coronary artery stenosis, but also an effective indicator for assessing early improvement in cardiac function in patients with CHD after PCI. In our study, echocardiograph was performed on the 4^th^ day after PCI, revealing a decrease in E/e’, and increases in GLS and LVEF-1. While PCI can immediately improve myocardial ischemia, the early postoperative period did not show significant changes in the LVEF of these patients, suggesting that LVEF may not be a sensitive marker for reflecting the recovery of myocardial ischemia. Further analysis demonstrated that a higher Gensini score and a larger stent diameter were associated with a positive impact on the improvement of LVEF-1. A higher Gensini score indicates more severe CHD, while a larger stent diameter suggests that a larger area of myocardial blood supply is affected. These findings indicate that patients with more severe coronary artery stenosis show more improvement in cardiac dysfunction evaluated by LVEF-1 after PCI.

Although we conducted a cross-sectional study on the relationship between LVEF-1 and coronary artery stenosis in patients with CHD, this study still has some limitations. Firstly, it is a small-scale study with a limited sample size, and there are some differences in the acquisition of ultrasound images and data measurements among different centers. Therefore, we need more multi-center data to support our conclusions. Additionally, we did not perform functional tests on the coronary arteries of these patients, nor did we investigate the relationship between LVEF-1 and coronary microcirculatory ischemia. Finally, cardiovascular drugs definitely have a certain impact on LVEF-1 although there have not been clearly reports. But all our selected patients received standardized drug treatment before CAG and during hospitalization, with no significant differences among these groups, which can minimize bias of our results.

## Conclusion

LVEF-1 is a valuable clinical indicator for assessing cardiac dysfunction caused by coronary artery stenosis. A decrease in LVEF-1 is strongly associated with the severity of CHD. An increase in LVEF-1 indicates the therapeutic effects of PCI, with a more significant improvement in LVEF-1 post-PCI in cases of more severe coronary artery stenosis.

## Consent for publication

Not applicable.

## Authors’ contributions

Study design: XSG, CT; Data collection and analysis: CT, BY, LX; Manuscript preparation: LX, YYZ, JXR. Data analysis: CT, LX. Coronary angiograph: XSG, XL, Echocardiograph: YYZ, JXR, BY. All the authors have approved the manuscript for publication.

## Data availability

The datasets used and/or analyzed during the current study are available from the corresponding author on reasonable request.

## Acknowledgments

We would like to thank Peng Hao from department of epidemiology, school of public health, Soochow University for help for data statistical analysis.

